# Clinically Recognised Alcohol Use Disorder and Risk of Dementia Subtypes in 1.3 Million Women: A Prospective Cohort Study

**DOI:** 10.64898/2026.09.23.26363779

**Authors:** Nasri G Fatih, David Hippisley-Cox, Klaus P Ebmeier, Joel Gelernter, Gillian K. Reeves, Sarah Floud, Maria D. Christodoulou, Anya Topiwala

**Author notes:** **Corresponding author:** Nasri Gabriel Fatih, Nuffield Department of Population Health, Big Data Institute,□University of Oxford,□Oxford, UK.

## Abstract

**Background:** Alcohol use disorder (AUD) is associated with increased dementia risk, but its long-term relationship with dementia, variation by age and subtype, and how this risk compares with that associated with alcohol consumption remain poorly characterised. We investigated these questions in a large prospective cohort of UK women.

**Methods:** We analysed Million Women Study participants recruited aged 50–64 years and followed for more than 20 years through linked health records. Women with hospital-recorded AUD were matched to up to five controls. Cox models estimated associations between time-varying AUD and incident dementia overall, by subtype, attained age, with exposure lags of up to 15 years and competing-risk analyses. Self-reported alcohol consumption was analysed separately.

**Findings:** Among 1,288,359 women, 20,070 with AUD were matched to 100,339 controls. Dementia occurred in 3,774 women with AUD and 8,239 matched controls. AUD was associated with all-cause dementia (HR=3·73[95% CI 3·52–3·95]), with strongest associations for mixed (5·03[3·88–6·51]), unspecified (4·29[3·92–4·70]), and vascular dementia (3·96[3·44–4·56]). Associations were substantially stronger before age 70 years (10·68[9·02–12·63]) than at ages 70-79 (3·75[3·47–4·04]) or ≥80 years (2·34[2·11–2·60]). Risk remained elevated with a 15-year lag (2·16[1·81–2·58]) and after accounting for competing mortality. Associations with self-reported alcohol consumption were substantially weaker.

**Interpretation:** Clinically recognised AUD identifies women at markedly increased and prolonged dementia risk, particularly at younger ages and for vascular and mixed dementia, not captured by drinking quantity alone. People with AUD should be considered a priority population for earlier dementia prevention and risk reduction.

**Funding:** Wellcome Trust Career Development Award to AT (306069/Z/23/Z); NIH grants P50AA012870 and R01DA054869 (JG). The Million Women Study is supported by the UK Medical Research Council (grant number UKRI1464)

## Introduction

Alcohol use disorder (AUD) is among the strongest potentially modifiable risk factors for dementia,^1^ with observational findings increasingly supported by genetic evidence implicating harmful alcohol use in its aetiology.^2^ However, important uncertainties remain. The 2026 WHO guidelines on risk reduction for cognitive decline and dementia highlight limited evidence in younger populations and individuals with AUD, a lack of large longitudinal studies to inform screening and prevention, and uncertainty regarding the mechanisms linking alcohol and dementia.^3^ Previous large-scale studies have not established whether excess dementia risk persists when AUD precedes dementia by many years, leaving uncertainty about reverse causation and diagnostic proximity. ^1^

Evidence on variation by age and dementia subtype is also limited, particularly among women, despite their substantial contribution to the global dementia burden.^4^ Dementia comprises a heterogeneous group of disorders with overlapping but distinct underlying pathophysiological mechanisms, and associations with AUD may differ across subtypes. AUD is strongly associated with hypertension, stroke, and atrial fibrillation, ^5,6^ and other vascular risk factors that could preferentially increase vascular dementia risk, whereas alcohol-related neurotoxicity, neuroinflammation, oxidative stress, and impaired amyloid clearance may contribute to neurodegenerative dementias.^7,8^ Associations may also differ across age.^1,9^ Stronger relative associations at younger ages could indicate a greater contribution of AUD to dementia risk, whereas at older ages dementia increasingly reflects multiple age-related pathologies and competing mortality becomes more important.

Interpretation further complicated by how alcohol exposure is measured. Most prospective studies rely on self-reported alcohol consumption assessed at a single time point, which may poorly capture longer-term drinking patterns ^10–12^ and is susceptible to under-reporting,^13^ and changes in consumption due to deteriorating health/ ^10^ Clinically recognised AUD captures a distinct, more severe dimension of alcohol-related harm, and when ascertained longitudinally, allows exposure status to change as AUD is recognised during follow-up. Directly comparing clinically recognised AUD with self-reported alcohol consumption within the same cohort can therefore establish whether drinking quantity alone captures the excess dementia risk associated with severe alcohol-related harm.

Temporality remains a further challenge. Previous studies have had limited ability to determine whether elevated dementia risk is concentrated around clinical recognition of AUD or persists when AUD is recorded many years before dementia diagnosis. Associations occurring predominantly over short intervals could partly reflect reverse causation, diagnostic proximity, or increased healthcare contact. Existing evidence is further limited by the predominance of studies in older populations, ^15,16^ relatively short follow-up ^1,15^ and insufficient power to examine less common dementia subtypes. We therefore examined associations between time-varying AUD and incident all-cause dementia and dementia subtypes among more than 1.3 million women in the Million Women Study for over two decades. We investigated variation across attained age, examined progressively longer intervals of up to 15 years between first recorded AUD and dementia, assessed the influence of competing mortality, and compared associations f with those observed for baseline self-reported alcohol consumption.

## Methods

### Study design and participants

Participants were drawn from the Million Women Study, a population-based prospective cohort of UK women aged 50–64 years recruited between May 1, 1996, and December 31, 2001, through the UK National Health Service breast screening programme.^17,18^ Participants completed questionnaires at recruitment and at four subsequent re-surveys, approximately 3, 8, 12, and 15 years later, covering sociodemographic factors, lifestyle, anthropometry, reproductive history, and health. Participants were linked to routinely collected NHS hospital and mortality records, including Hospital Episode Statistics in England and Scottish Morbidity Records in Scotland. ^17,18^

### Exposure

AUD was defined from hospital records using International Classification of Diseases, 10th Revision (ICD-10) codes for AUD or alcohol-related harm. Alcohol-related dementia codes were excluded from the exposure definition because they formed part of the dementia outcome classification. The date of the first recorded AUD diagnosis defined exposure onset, which was modelled as a time-varying variable. The code list was developed and reviewed with specialist clinical input and is provided in the supplementary material and in machine-readable format in the study GitHub repository (https://github.com/Topiwala-Lab/mws-aud-dementia).

Self-reported alcohol consumption at recruitment, measured in UK units per week, was analysed separately as a secondary exposure. Self-reported alcohol consumption was not used to define hospital-recorded AUD exposure. Further details are provided in the Supplementary Methods.

### Outcomes

Dementia cases were identified from linked hospital records using ICD-10 codes. Outcomes included all-cause dementia and Alzheimer’s disease, vascular, mixed, unspecified, Parkinson’s disease dementia/Lewy body dementia, and frontotemporal dementia.

Dementia subtypes were assigned from all available ICD-10-coded records using a prespecified algorithm developed with specialist clinical input. Women with one specific dementia subtype recorded were assigned to that subtype; those with two or more specific subtypes were classified as having mixed dementia. An unspecified dementia diagnosis was superseded by a specific subtype if one was subsequently recorded. The event date was the earliest qualifying dementia diagnosis, irrespective of later subtype refinement.

Mild cognitive impairment was identified separately and was not classified as dementia. Women with mild cognitive impairment followed by a qualifying dementia diagnosis were assigned the subsequent dementia subtype, with the first qualifying dementia diagnosis used as the event date. Women with mild cognitive impairment only remained in the primary risk set and were excluded in a sensitivity analysis.

Alcohol-related dementia codes contributed to the all-cause dementia outcome but were not included among the primary subtype-specific analyses because of their diagnostic and conceptual overlap with alcohol-related harm. Dementia-related ICD-10 code lists are provided in the supplementary material, with machine-readable code lists, analysis scripts, and supporting results available in the study GitHub repository (https://github.com/Topiwala-Lab/mws-aud-dementia).

### Confounders

Information on education, smoking status, treatment for depression or anxiety, diabetes, and cardiovascular disease was obtained from questionnaire data. Education was modelled categorically as tertiary qualifications, secondary qualifications, technical qualifications, no qualifications but completed compulsory schooling, did not complete compulsory school/no school, or missing/unknown. Smoking status was modelled as never smoker, past smoker, current smoker, or not current/unknown current status.

Area deprivation was measured using the Townsend deprivation index. Participants were assigned to a census small area using their residential postcode and given the corresponding Townsend score for that area. Deprivation was modelled in quintiles from quintile 1, least deprived, to quintile 5, most deprived, with missing/unknown deprivation retained as a separate category. Region was defined according to geographical region of residence at recruitment. Date of death was obtained through linkage to national mortality records.

### Statistical analysis

#### Analyses were conducted using R version 4.3.2

Baseline characteristics were summarised using medians and interquartile ranges for continuous variables and counts and percentages for categorical variables. Each participant with AUD was matched to up to five controls without replacement. Controls were required to have the same education, smoking-status, and area-deprivation categories and were then selected by nearest-neighbour matching on age at recruitment within a 2-year caliper.

Covariate balance was assessed using standardised mean differences.

We fitted matched Cox proportional hazards models with attained age as the underlying time scale and AUD modelled as a time-varying exposure. Participants contributed unexposed person-time until their first recorded AUD diagnosis and exposed person-time thereafter. Model 1 adjusted for education, smoking status, area deprivation, and region, with stratification by matched set. Model 2 additionally adjusted for treatment for depression or anxiety, diabetes, and cardiovascular disease.

Proportionality was assessed using Schoenfeld residuals^19^, recognising that proportional hazards need not be treated as a binary requirement for survival analysis^20^. Since associations between AUD and dementia varied by attained age, age-specific hazard ratios were estimated within prespecified attained-age bands of <70, 70–79, and ≥80 years. Proportionality was subsequently re-assessed after allowing the AUD association to vary across these age bands. For outcomes with residual evidence of non-proportionality, supplementary models allowed the association between AUD and dementia to vary continuously with attained age using an interaction between AUD and a natural cubic spline for attained age, with three interior knots at the 25th, 50th, and 75th percentiles of event age. These analyses were used to assess whether the age-specific associations reflected a smooth change with increasing age rather than being dependent on the selected age-band cut-points.

To characterise how the magnitude of the association varied according to the temporal relationship between first recorded AUD and subsequent dementia, we examined the AUD– dementia association across annual exposure lags from 0 to 15 years, with the 0-year estimate corresponding to the primary time-varying exposure model. For each lag greater than 0 years, participants remained classified as unexposed until the specified interval had elapsed after their first recorded AUD record, after which they were classified as exposed. Dementia events occurring during the lag period were retained as events during unexposed person-time.

Self-reported alcohol consumption at recruitment was analysed separately in the full model-eligible cohort without matching. Participants with missing alcohol consumption data, prevalent dementia, or dementia diagnosed within 12 months of recruitment were excluded from these analyses. Alcohol consumption was categorised as 0, <1, 1–6, 7–13, 14–20, and ≥21 units/week, with 1–6 units/week as the reference category. Cox models used attained age as the underlying time scale. Model 1 adjusted for education, smoking status, area deprivation, and region, and Model 2 additionally adjusted for treatment for depression or anxiety, diabetes, and cardiovascular disease.

Missingness was assessed using prespecified quality-control checks that quantified the number and proportion of missing observations for each model variable and tracked exclusions through model eligibility. Participants with missing data for covariates required by a given model were excluded from that analysis. Participants with missing self-reported alcohol consumption were additionally excluded from the baseline alcohol-consumption analyses.

Since death precludes a subsequent diagnosis of dementia, we additionally fitted Fine–Gray subdistribution hazard models to estimate associations with the cumulative incidence of dementia in the presence of competing mortality^21^. These analyses complemented the primary cause-specific Cox models, which estimated the cause-specific hazard of dementia among individuals who remained alive and dementia-free. We also fitted a cause-specific Cox model for death before dementia. Absolute cumulative incidences of dementia and death before dementia were estimated using the Aalen–Johansen estimator, with dementia and death treated as competing events, and were summarised at 5 and 10 years^22^.

In supplementary analyses, baseline alcohol consumption associations were additionally estimated within attained-age bands. To assess whether prodromal cognitive impairment without a recorded dementia diagnosis influenced the findings, analyses were repeated after excluding participants with mild cognitive impairment only. To account for secular changes in hospital coding, diagnostic practice, and dementia ascertainment over the study period, follow-up was split by calendar period, and calendar period was included as a time-varying covariate. Further implementation details are provided in the Supplementary Methods.

## Results

Among 1,288,359 eligible women, 20,070 (1·6%) had a hospital-recorded AUD diagnosis during follow-up. Mean follow-up was 21·96 years (SD 5·74), during which 119,304 women were diagnosed with dementia: 3,774 (18.8%) among women with a recorded AUD and 115,530 (9.1%) among women without recorded AUD. Before matching, women with AUD differed from those without AUD across several baseline characteristics, including smoking status, area deprivation, self-reported alcohol consumption, and treatment for depression or anxiety (Supplementary Table 1). The matched cohort comprised 20,070 women with AUD and 100,339 controls. Matching substantially improved covariate balance, and mean age at recruitment was 55·88 years in both groups (Supplementary Table 2). Alcohol-related dementia contributed to the all-cause dementia outcome but was considered separately from the primary subtype-specific analyses because of its diagnostic overlap with alcohol-related harm.

In the matched cohort, 3,774 dementia cases occurred among women who had a recorded AUD diagnosis at some point during follow-up and 8,239 among matched controls. In the time-varying analysis, 2,758 dementia events occurred during AUD-exposed person-time and 9,198 during unexposed person-time. AUD was associated with a substantially higher cause-specific hazard of all-cause dementia (Model 2 HR 3·73, 95% CI 3·52–3·95; Figure 1).

**Figure 1.**
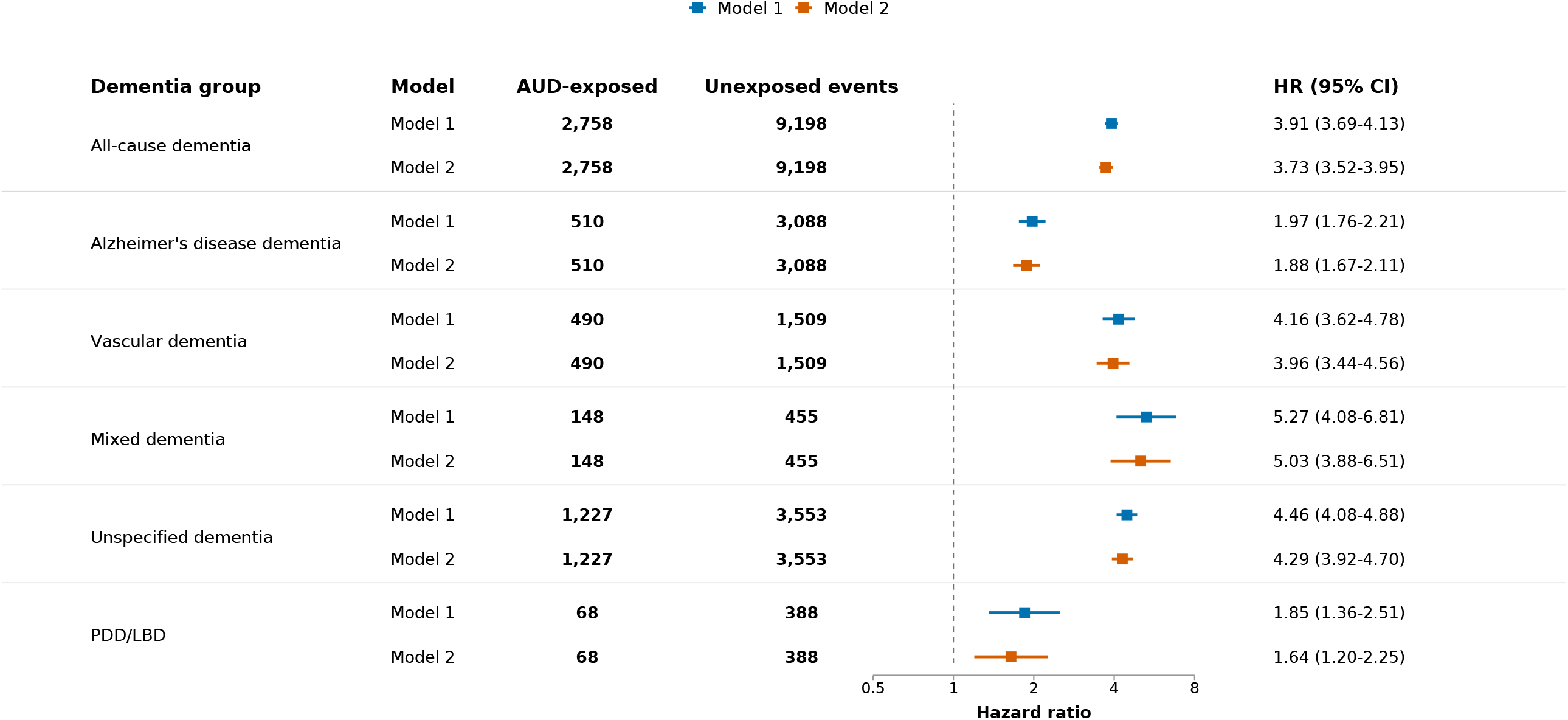
Associations between time-varying alcohol use disorder and incident dementia. Hazard ratios (HRs) and 95% confidence intervals are shown for the association between time-varying alcohol use disorder (AUD) and all-cause dementia and dementia subtypes in the matched cohort (N=120,409; 20,070 women with AUD and 100,339 matched controls). Participants contributed unexposed person-time until their first recorded AUD diagnosis and exposed person-time thereafter. Model 1 adjusted for education, smoking status, area deprivation, and region, with stratification by matched set. Model 2 additionally adjusted for treatment for depression/anxiety, diabetes, and cardiovascular disease. AUD and No-AUD event counts denote dementia events occurring during AUD-exposed and unexposed person-time, respectively. The vertical reference line indicates HR=1. PDD/LBD=Parkinson’s disease dementia/Lewy body dementia.

Associations were observed across the major dementia subtypes but varied considerably in magnitude. The strongest associations were observed for mixed dementia (HR 5·03, 95% CI 3·88–6·51), unspecified dementia (4·29, 3·92–4·70), and vascular dementia (3·96, 3·44–4·56). Associations were smaller for Alzheimer’s disease (1·88, 1·67–2·11) and Parkinson’s disease dementia/Lewy body dementia (1·64, 1·20–2·25). Estimates were only modestly attenuated after additional adjustment in Model 2. Frontotemporal dementia was uncommon, with 64 events overall and 12 events occurring during AUD-exposed person-time, limiting reliable subtype-specific inference.

The association between AUD and all-cause dementia varied substantially across attained age (Figure 2). The Model 2 HR was 10·68 (95% CI 9·02–12·63) at ages <70 years, 3·75 (3·47– 4·04) at ages 70–79 years, and 2·34 (2·11–2·60) at ages ≥80 years. A similar pattern of stronger relative associations at younger ages was evident for several dementia subtypes. For unspecified dementia, HRs were 10·75 (8·19–14·11), 4·94 (4·37–5·58), and 2·43 (2·09–2·84) across the same age groups, respectively; for mixed dementia, corresponding HRs were 12·70 (6·16–26·18), 4·61 (3·36–6·33), and 3·15 (1·72–5·76). Although relative associations were strongest at younger ages, most dementia events occurring during AUD-exposed person-time occurred at older ages: 491 at ages <70 years, 1,471 at ages 70–79 years, and 796 at ages ≥80 years. The incidence rate of all-cause dementia at ages <70 years was 8·97 per 1,000 person-years during AUD-exposed person-time compared with 0·60 during unexposed person-time. At ages ≥80 years, corresponding rates were 61·59 and 26·08 per 1,000 person-years. Supplementary continuous age-varying models showed the same overall decline in the relative association with increasing attained age.

**Figure 2.**
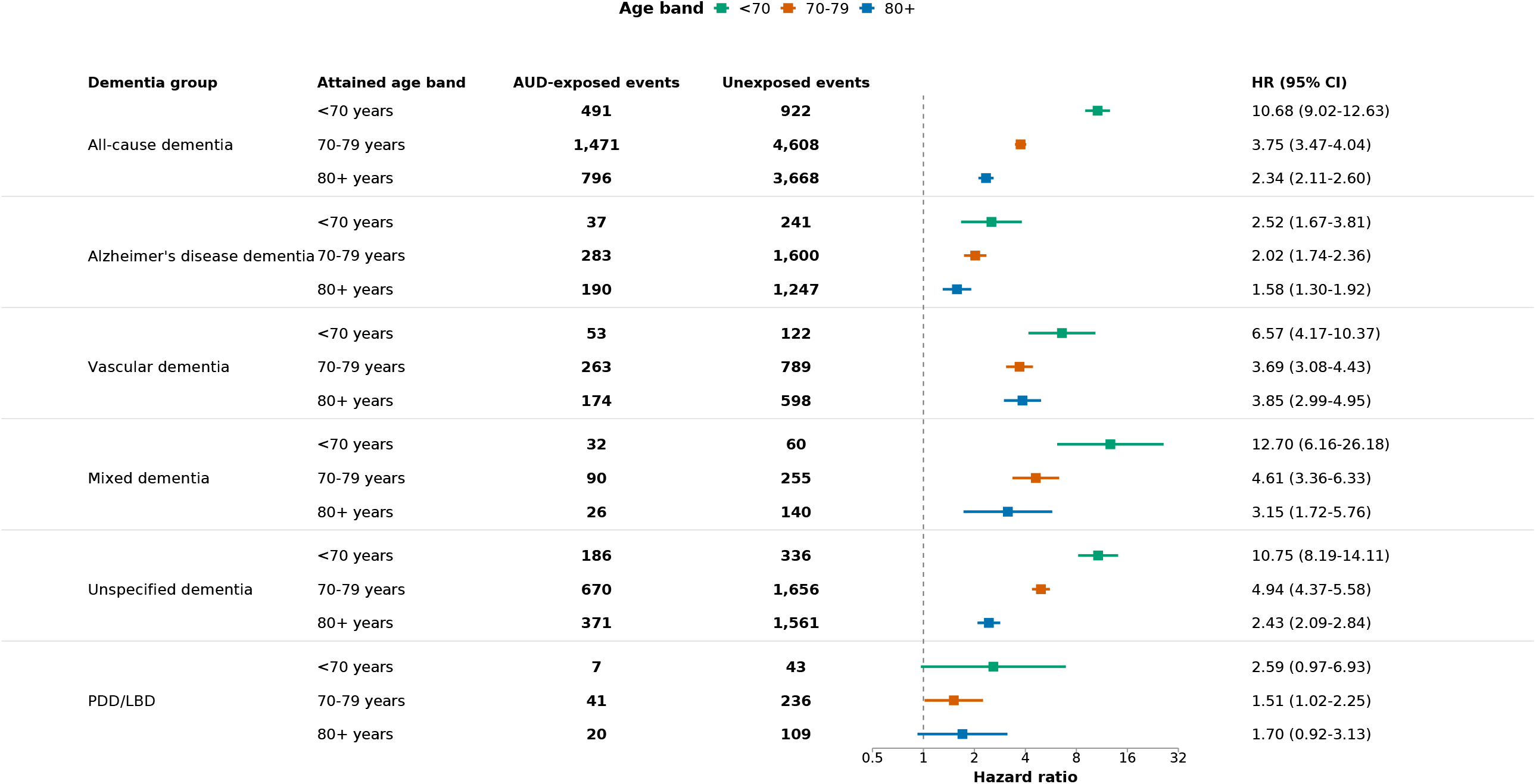
Association between time-varying alcohol use disorder and incident dementia by attained age. Hazard ratios (HRs) and 95% confidence intervals are shown for the association between time-varying alcohol use disorder (AUD) and all-cause dementia and dementia subtypes within attained-age bands of <70, 70–79, and ≥80 years. Estimates are from Model 2, adjusted for education, smoking status, area deprivation, region, treatment for depression/anxiety, diabetes, and cardiovascular disease, with stratification by matched set. AUD and No-AUD event counts denote dementia events occurring during AUD-exposed and unexposed person-time within each attained-age band, respectively. The vertical reference line indicates HR=1. PDD/LBD=Parkinson’s disease dementia/Lewy body dementia.

Associations generally attenuated with increasing temporal separation between first recorded AUD and dementia diagnosis (Figure 3). For all-cause dementia, the Model 2 HR declined from 3·73 (95% CI 3·52–3·95) with no exposure lag to 3·11 (2·90–3·34) with a 3-year lag, 2·82 (2·60–3·06) with a 5-year lag, 2·56 (2·28–2·88) with a 10-year lag, and 2·16 (1·81–2·58) with a 15-year lag. Associations therefore remained elevated even when AUD exposure classification was delayed by 10–15 years. Associations for Alzheimer’s disease, vascular, mixed, and unspecified dementia also generally attenuated over longer lags, although estimates became less precise at longer intervals, particularly for less common subtypes.

**Figure 3.**
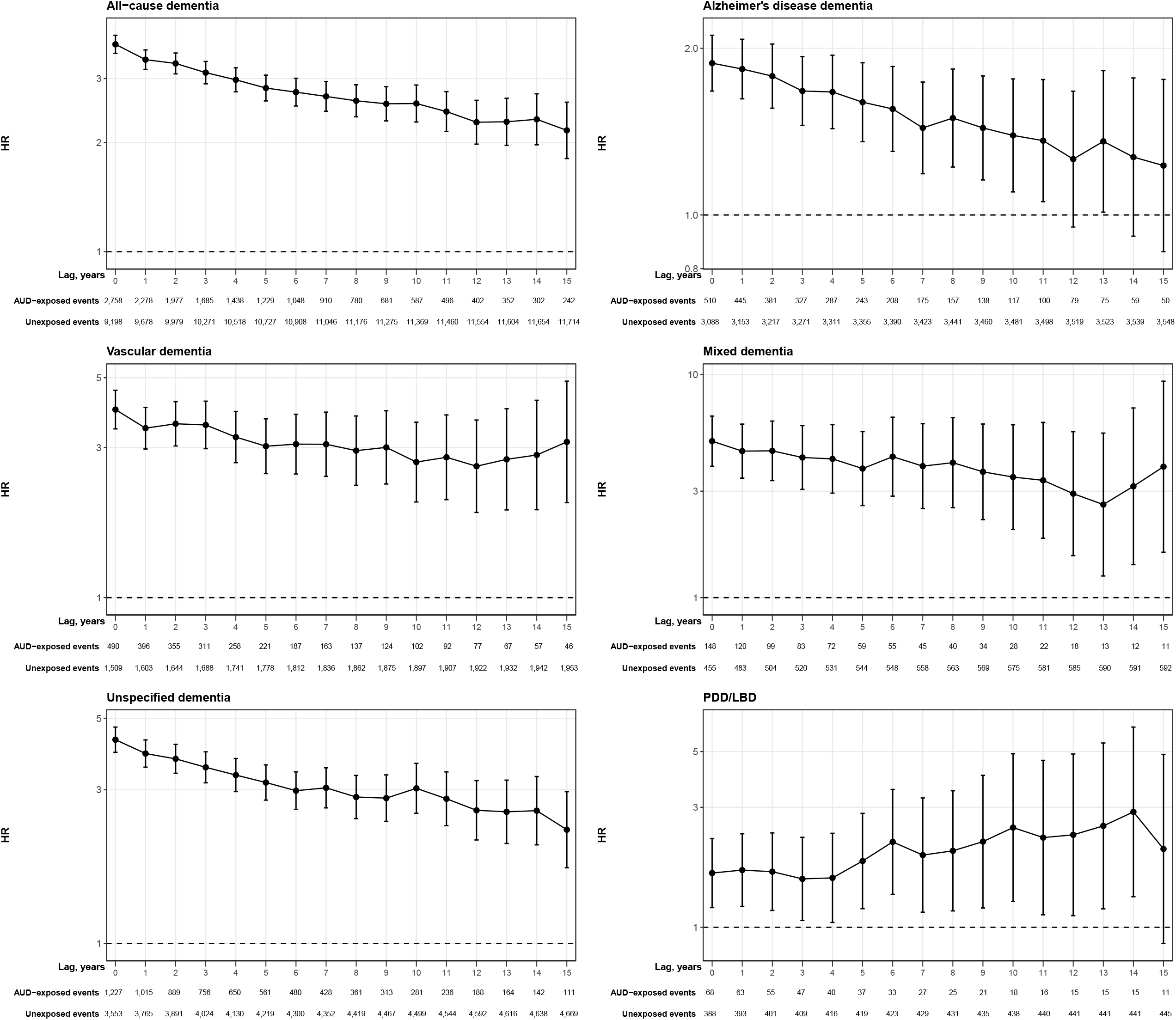
Association between alcohol use disorder and incident dementia across increasing exposure lags. Hazard ratios (HRs) and 95% confidence intervals (CIs) are shown for the association between alcohol use disorder (AUD) and all-cause dementia, Alzheimer’s disease dementia, vascular dementia, mixed dementia, unspecified dementia, and Parkinson’s disease dementia/Lewy body dementia across annual exposure lags from 0 to 15 years. The 0-year lag corresponds to the primary time-varying exposure model. For each lag of *L* years, participants remained classified as unexposed until *L* years after their first recorded AUD diagnosis and were classified as exposed thereafter. Dementia events occurring during the lag period were retained as events during unexposed person-time rather than excluded. Estimates are from Model 2, adjusted for education, smoking status, area deprivation, region, treated depression/anxiety, diabetes, and cardiovascular disease, with stratification by matched set. Event-count columns show the numbers of “AUD-exposed events” and “unexposed events” at each lag. These analyses characterise how the AUD– dementia association changes with increasing temporal separation between first recorded AUD and dementia diagnosis. Y-axis scales differ where indicated, including for the 15-year lag, and should therefore be interpreted within each panel. The horizontal reference line indicates HR=1.

AUD was also strongly associated with death before dementia (Model 2 cause-specific HR 3·59, 95% CI 3·48–3·71; Figure 4). In Fine–Gray analyses accounting for death as a competing event, AUD remained associated with a higher cumulative incidence of all-cause dementia, although the magnitude of the association was smaller than in the primary cause-specific Cox model (subdistribution HR 2·35, 95% CI 2·25–2·46). Corresponding subdistribution HRs were 2·77 (2·27–3·38) for mixed dementia, 2·69 (2·51–2·88) for unspecified dementia, 2·33 (2·09–2·59) for vascular dementia, 1·20 (1·09–1·32) for Alzheimer’s disease, and 1·29 (0·99–1·68) for Parkinson’s disease dementia/Lewy body dementia. At 10 years, the cumulative incidence of dementia was 14·1% among women with AUD compared with 4·9% among matched controls, an absolute difference of 9·2 percentage points. The corresponding cumulative incidence of death before dementia was 41·6% among women with AUD and 15·4% among controls, an absolute difference of 26·2 percentage points. At 5 years, cumulative dementia incidence was 9·2% versus 2·2%, and cumulative mortality was 27·6% versus 7·1% (Supplementary Table 10).

**Figure 4.**
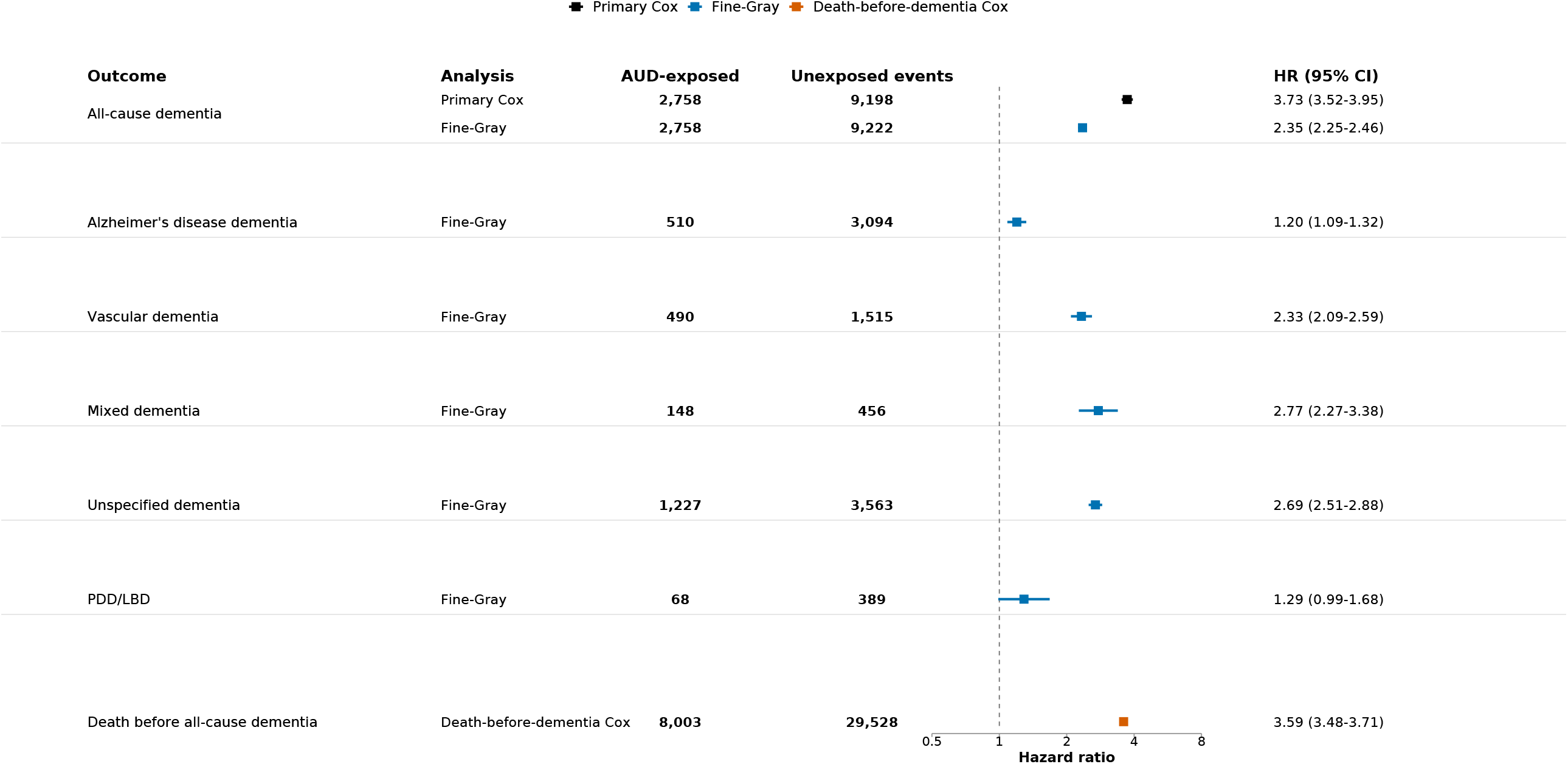
Alcohol use disorder, dementia, and competing mortality. Associations between time-varying alcohol use disorder (AUD), dementia, and death before dementia are shown using cause-specific Cox and Fine–Gray subdistribution hazard models. Cause-specific Cox models estimate the hazard of dementia among participants who remain alive and dementia-free, whereas Fine–Gray models estimate associations with the cumulative incidence of dementia in the presence of death as a competing event. A separate cause-specific Cox model estimates the association between AUD and death before dementia. Estimates are from Model 2, adjusted for education, smoking status, area deprivation, region, treatment for depression/anxiety, diabetes, and cardiovascular disease, with stratification by matched set where applicable. For dementia analyses, AUD and No-AUD event counts denote dementia events occurring during exposed and unexposed person-time; for the mortality analysis, they denote deaths before dementia occurring during exposed and unexposed person-time. HR=hazard ratio; sHR=subdistribution hazard ratio; PDD/LBD=Parkinson’s disease dementia/Lewy body dementia.

Associations between baseline self-reported alcohol consumption and dementia were substantially weaker than those observed for hospital-recorded AUD (Figure 5). Compared with women reporting 1–6 units/week, associations showed a broadly non-linear pattern, with higher dementia rates among women reporting no alcohol consumption and more modest increases in some higher-consumption categories. These associations also varied by attained age. For all-cause dementia at ages <70 years, HRs were 1·31 for 0 units/week, 1·06 for <1 unit/week, 1·09 for 7–13 units/week, 1·27 for 14–20 units/week, and 1·51 for ≥21 units/week, relative to 1–6 units/week. Differences were smaller at older ages. A similar pattern was evident for unspecified dementia, for which the HR at ages <70 years was 1·76 among women reporting ≥21 units/week. Full estimates are provided in the Supplementary Tables.

**Figure 5.**
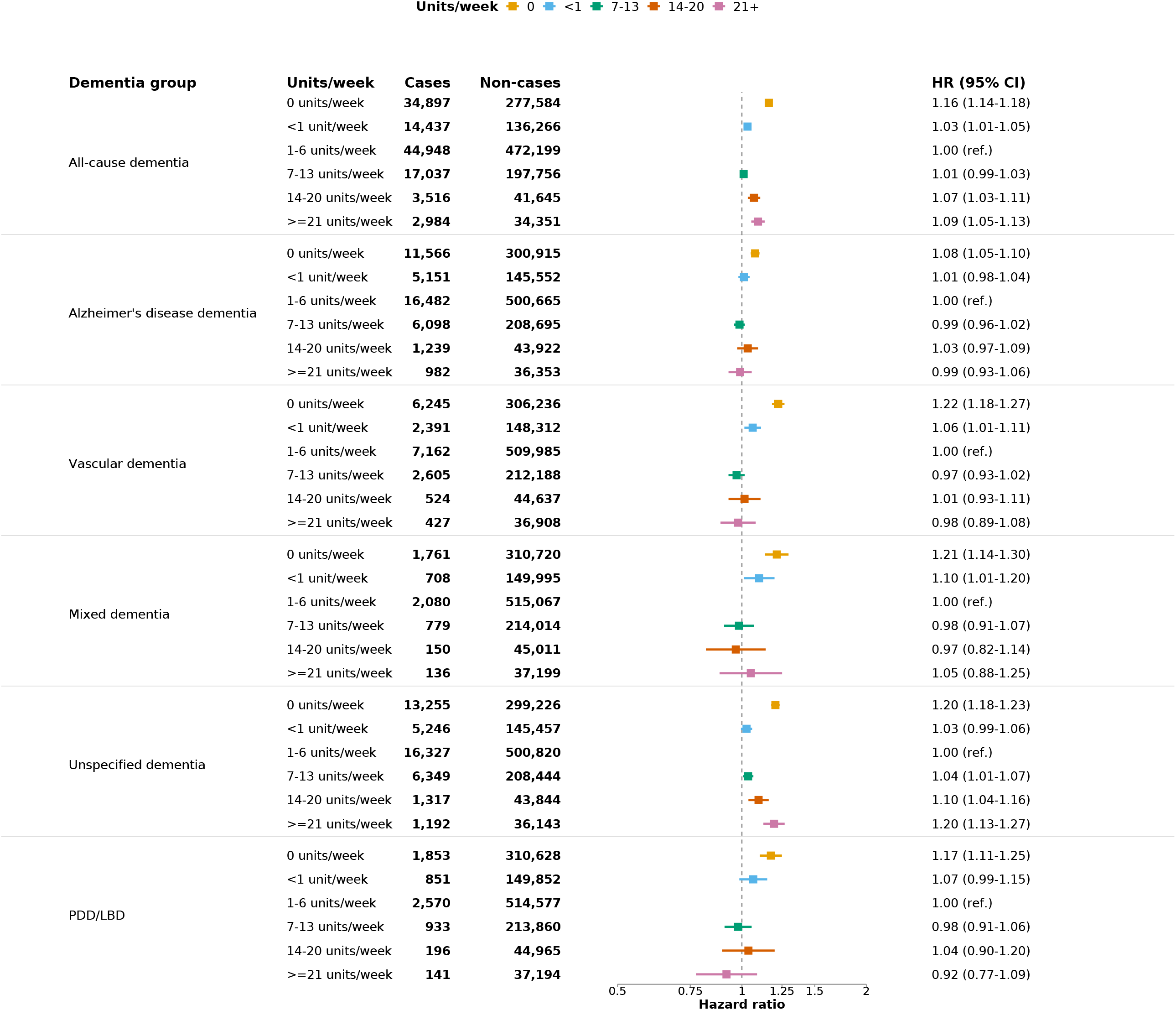
Association between baseline self-reported alcohol consumption and incident dementia. Hazard ratios (HRs) and 95% confidence intervals are shown for associations between self-reported alcohol consumption at recruitment and all-cause dementia and dementia subtypes in the full model-eligible cohort. Alcohol consumption was categorised as 0, <1, 1–6, 7–13, 14–20, and ≥21 UK units/week, with 1–6 units/week as the reference category. Models used attained age as the underlying time scale and were not matched or stratified by matched set. Estimates are from Model 2, adjusted for education, smoking status, area deprivation, region, treatment for depression/anxiety, diabetes, and cardiovascular disease. Participants with prevalent dementia, dementia diagnosed within 12 months of recruitment, or missing baseline alcohol-consumption data were excluded. Cases and non-cases denote the number of women with and without the relevant dementia outcome within each baseline alcohol-consumption category. The vertical reference line indicates HR=1. PDD/LBD=Parkinson’s disease dementia/Lewy body dementia.

Results were broadly unchanged after excluding women with mild cognitive impairment without a recorded dementia diagnosis. After exclusion of 547 women with mild-cognitive-impairment-only records and removal of matched sets rendered incomplete by this exclusion, 118,880 women remained in the analysis. Adjustment for calendar period as a time-varying covariate likewise produced estimates similar to those from the primary analyses.

## Discussion

In this large prospective cohort of UK women followed for more than two decades, clinically recognised AUD was associated with substantially higher rates of all-cause dementia, with marked variation by dementia subtype and age. Associations were strongest for mixed, unspecified, and vascular dementia and were particularly pronounced at younger attained ages. Although, associations attenuated with increasing time since first recorded AUD, they remained elevated after15 years, and despite substantial competing mortality. In contrast, baseline self-reported alcohol consumption showed considerably weaker associations, suggesting that clinically recognised AUD identifies a high-risk population not captured by drinking quantity alone.

The particularly strong associations with vascular, mixed, and unspecified dementia are consistent with the possibility that vascular and multisystem pathways contribute substantially to dementia risk associated with AUD. AUD is associated with hypertension, stroke, atrial fibrillation, and other vascular disease, while harmful alcohol use may also contribute to neurotoxicity, neuroinflammation, oxidative stress, nutritional deficiencies, and metabolic dysfunction.^5–8^ The comparatively weaker association with Alzheimer’s disease is consistent with previous studies in which AUD has generally shown stronger associations with vascular and other forms of dementia than with Alzheimer’s disease. ^1,23^. These findings should not, however, be interpreted as demonstrating subtype-specific causal mechanisms. Dementia diagnoses in routine clinical data are imperfect proxies for underlying neuropathology, and mixed pathologies are common^24,25^ (Nichols et al., 2023; Schneider et al., 2007). Residual confounding by vascular, socioeconomic, behavioural, and other factors also remains possible.

The marked variation in relative associations across attained age is also important. AUD was associated with an approximately ten-fold higher hazard of all-cause dementia before age 70 years, compared with a little over two-fold higher hazard at ages 80 years and older. Stronger relative associations at younger ages may indicate that severe alcohol-related harm represents a larger proportion of the overall determinants of dementia at ages when background dementia incidence is otherwise low ^9^. At older ages, dementia increasingly reflects multiple age-related pathologies and competing causes of disease and death, which may reduce the relative association with any individual risk factor.^24,25^ Nevertheless, the lower relative association at older ages should not be interpreted as implying little absolute burden. Most dementia events occurring during AUD-exposed person-time arose after age 70 years, and absolute dementia incidence was substantially higher at older ages. The relative and absolute importance of AUD may therefore differ across the life course.

The progressive attenuation of associations with increasing exposure lag provides insight into the temporal relationship between clinically recognised AUD and subsequent dementia. Larger estimates at shorter intervals suggest that reverse causation, diagnostic proximity, or increased healthcare contact around the time of AUD recognition may contribute to the magnitude of the association.^1^ However, associations remained elevated when AUD exposure classification was delayed by 10–15 years, indicating these factors are unlikely to explain the association entirely and that clinically recognised AUD identifies women with elevated dementia risk over a prolonged period. Longer lags may also exclude dementia occurring relatively soon after alcohol-related harm, including cases in which alcohol may have contributed causally to cognitive impairment. ^26^ Thus, although the findings strengthen the temporal evidence linking AUD with subsequent dementia, they do not establish causality, and residual confounding and uncertainty about the true onset of harmful alcohol use remain. Competing mortality was substantial among women with AUD. Previous alcohol–dementia studies have similarly highlighted death as an important competing event, particularly in populations with severe alcohol-related harm.^23^ The smaller association observed in our Fine–Gray analyses than in the primary cause-specific Cox models is consistent with the high incidence of death before dementia in this group, which reduces the opportunity for dementia to be diagnosed. Nevertheless, the cumulative incidence of dementia remained substantially higher among women with AUD than among matched controls despite their markedly higher mortality. Thus, competing death attenuated the observed association but did not fully explain the excess dementia burden associated with AUD.

Associations with baseline self-reported alcohol consumption were substantially weaker than those observed for clinically recognised AUD, although these measures capture different aspects of alcohol exposure. Consumption was assessed at recruitment and may not reflect longer-term drinking patterns or subsequent changes in intake. ^10,11,14 10^ Moreover, relatively few women reported high consumption, and our highest category (>21 units/week) captures substantially lower exposure than is likely among many people with AUD, limiting our ability to compare the effects of heavy self-reported drinking directly with clinically recognised AUD. Self-reported alcohol intake may also be underestimated,^13^ while non-drinkers include former drinkers who may have stopped because of deteriorating health. The observed U-shaped association should therefore not be interpreted as evidence that low levels of alcohol consumption are protective against dementia. In contrast, time-varying AUD captures the emergence of a severe alcohol-related phenotype during follow-up. The considerably stronger association with dementia may therefore reflect both greater severity and longitudinally ascertainment. Future studies should incorporate clinically recognised AUD and longitudinal markers of alcohol-related harm, as reliance on a single baseline measure may fail to capture the dementia risk associated with severe alcohol exposure.

This study has several strengths. The Million Women Study provided a large prospective population with more than two decades of follow-up and sufficient a incident dementia cases to examine across age and dementia subtypes. Modelling AUD as a time-varying exposure preserved the temporal ordering of first recorded AUD and subsequent dementia and avoided classifying women as exposed on the basis of future diagnosis. Matching substantially improved measured baseline comparability between women with and without AUD, while use of attained age as the underlying time scale and explicit assessment of non-proportionality allowed age-related variation in associations to be characterised. Progression exposure lags of up to 15-years enable assessment of whether associations persisted well beyond the period following AUD recognition. Finally, competing risk analyses addressed the substantial excess mortality associated with AUD, while parallel analyses of self-reported alcohol consumption allowed two complementary dimensions of alcohol exposure to be examined within the same cohort.

Several limitations should also be considered. Hospital-recorded AUD is likely to identify more severe alcohol-related harm and will miss many individuals with harmful drinking or AUD managed outside hospital settings. Such exposure misclassification could attenuate associations and means that the findings should not be extrapolated directly to all levels of alcohol consumption or AUD severity. The date of first recorded AUD does not necessarily represent the biological onset of harmful alcohol use, which may have preceded clinical recognition by many years. Dementia subtype classification was based on routinely recorded clinical diagnoses rather than neuropathological confirmation, and diagnostic misclassification could reduce subtype specificity, particularly for mixed and unspecified dementia. Differential healthcare contact among women with AUD could also increase opportunities for dementia detection, particularly close to AUD recognition, although the persistence of associations over long exposure lags suggests that this is unlikely to explain the findings entirely. Residual and unmeasured confounding cannot be excluded despite matching and multivariable adjustment. The cohort included women recruited through the NHS breast screening programme at ages 50–64 years, and the findings may not generalise directly to men, younger populations, or populations with different healthcare systems or drinking patterns. Women entering the cohort may also have differed from the general population in health and socioeconomic characteristics. Some less common dementia subtypes had relatively few events, limiting precision. Alcohol-related dementia was not examined as a primary subtype-specific outcome because of its diagnostic and conceptual overlap with alcohol-related harm. Nevertheless, findings were broadly unchanged after excluding women with mild cognitive impairment only and after adjustment for calendar-period changes.

The cohort included women recruited through the NHS breast screening programme at ages 50–64 years, and the findings may not generalise directly to men, younger populations, or populations with different healthcare systems or drinking patterns. Women entering the cohort may also have differed from the general population in health and socioeconomic characteristics. Some less common dementia subtypes had relatively few events, limiting precision. Alcohol-related dementia was not examined as a primary subtype-specific outcome because of its diagnostic and conceptual overlap with alcohol-related harm. Nevertheless, findings were broadly unchanged after excluding women with mild cognitive impairment only and after adjustment for calendar-period changes.

In conclusion, clinically recognised AUD was associated with markedly elevated dementia risk in women, particularly at younger ages and for vascular, mixed, and unspecified dementia. Although associations attenuated with increasing time since first recorded AUD, excess risk persisted over10–15 years and despite substantial competing mortality. Clinically recognised AUD therefore identifies a population at prolonged elevated risk of dementia and a potential priority for earlier dementia prevention, including treatment of alcohol-related harm and identification and management of co-occurring vascular and metabolic risk. The substantially weaker associations with baseline self-reported alcohol consumption further suggest that drinking quantity alone does not capture the dementia risk associated with severe, alcohol-related harm.

## Data Availability

The Million Women Study data are not publicly available. Access to the data is governed by the Million Women Study data access and sharing policy. No new primary data were collected for this study. Analysis scripts are available from the study GitHub repository.

https://github.com/Topiwala-Lab/mws-aud-dementia

https://www.ceu.ox.ac.uk/research/the-million-women-study/data-access-and-sharing/data-access-policy

## Author contribution

NF and AT conceived the study. NF, AT, and MC developed the statistical analysis plan. NF prepared and harmonised the analytic dataset, conducted all analyses, produced the tables and figures, interpreted the findings, and wrote the first draft of the manuscript. NF, AT and MC contributed to the study design, interpretation of findings, and critical revision of the manuscript. JG, SF, GR, and KE, provided critical revision of the manuscript. All authors reviewed and approved the final version of the manuscript and accept responsibility for the decision to submit for publication.

## Data sharing

The Million Women Study data sharing policy can be accessed online: https://www.ceu.ox.ac.uk/research/the-million-women-study/data-access-and-sharing/data-access-policy. No new data were created during this study.

## Acknowledgements

The authors thank the women who have participated in the Million Women Study as well as the staff from the participating NHS breast screening centres. This work used data provided by patients and collected by the NHS as part of their care and support; the authors thank NHS England and Public Health Scotland for the health outcomes data.

## Ethics approval and consent

Ethical approval was provided by the East of England-Cambridge South Research Ethics Committee (REC 97/5/001). All participants gave consent for follow-up through medical records.

---

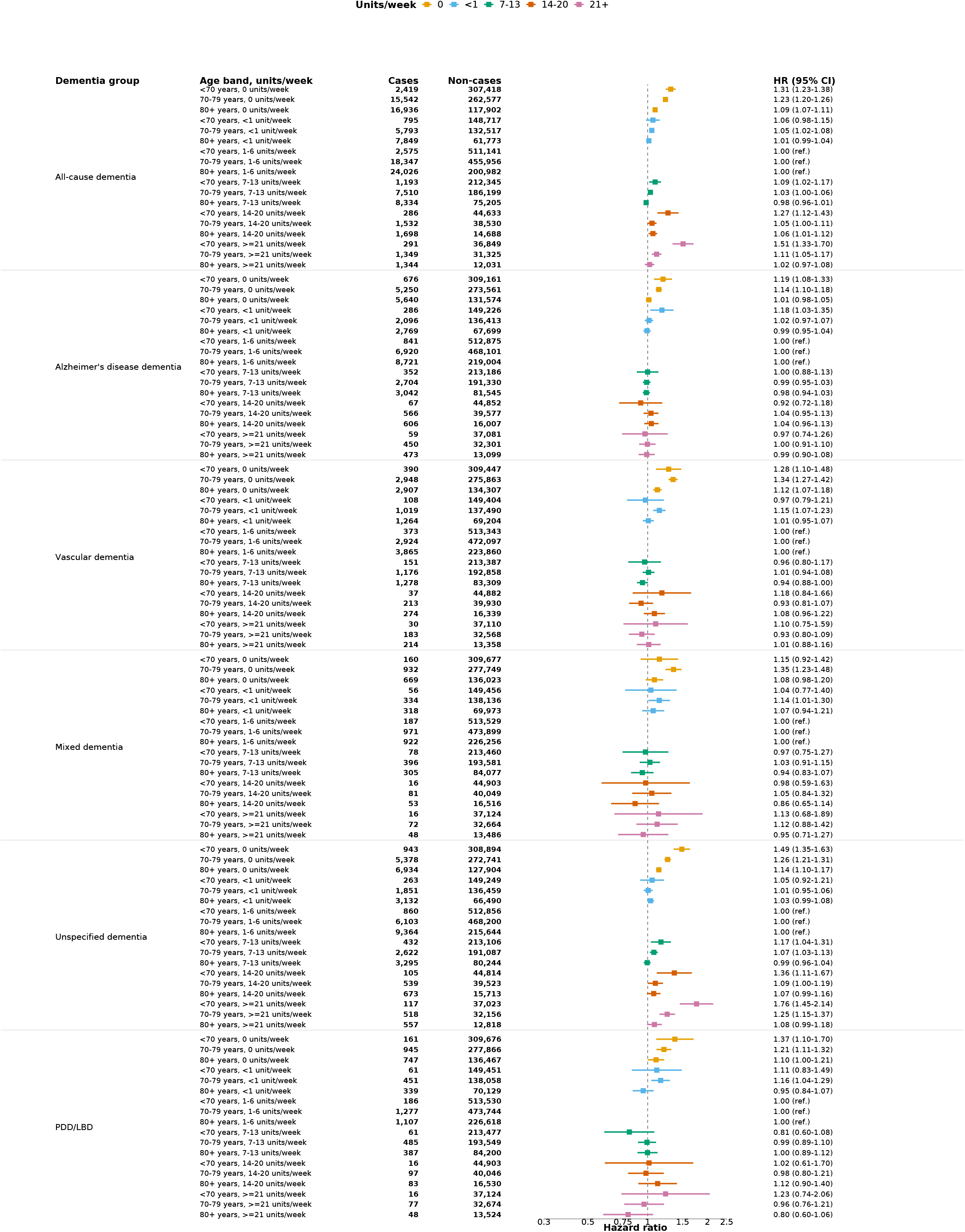

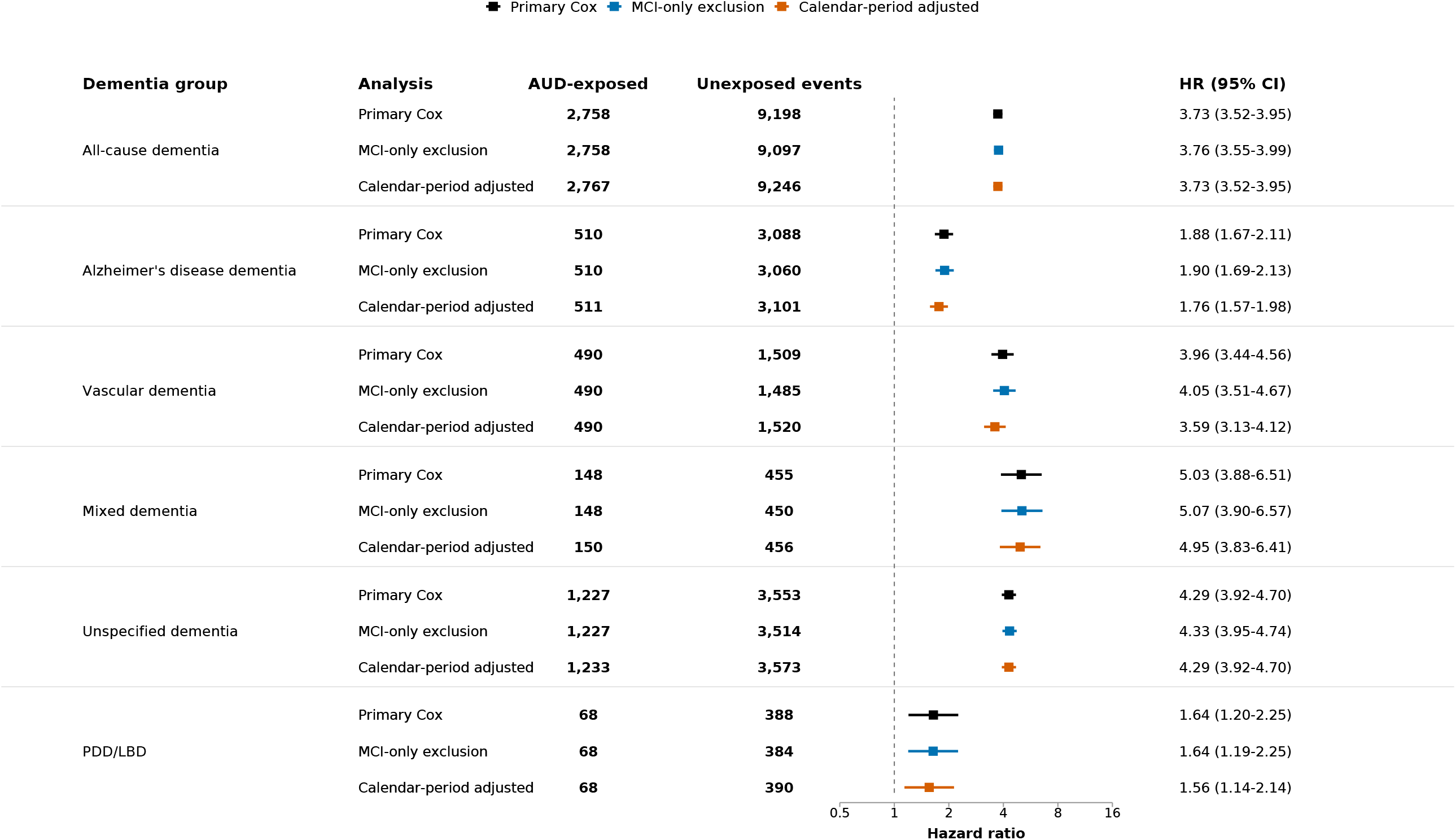

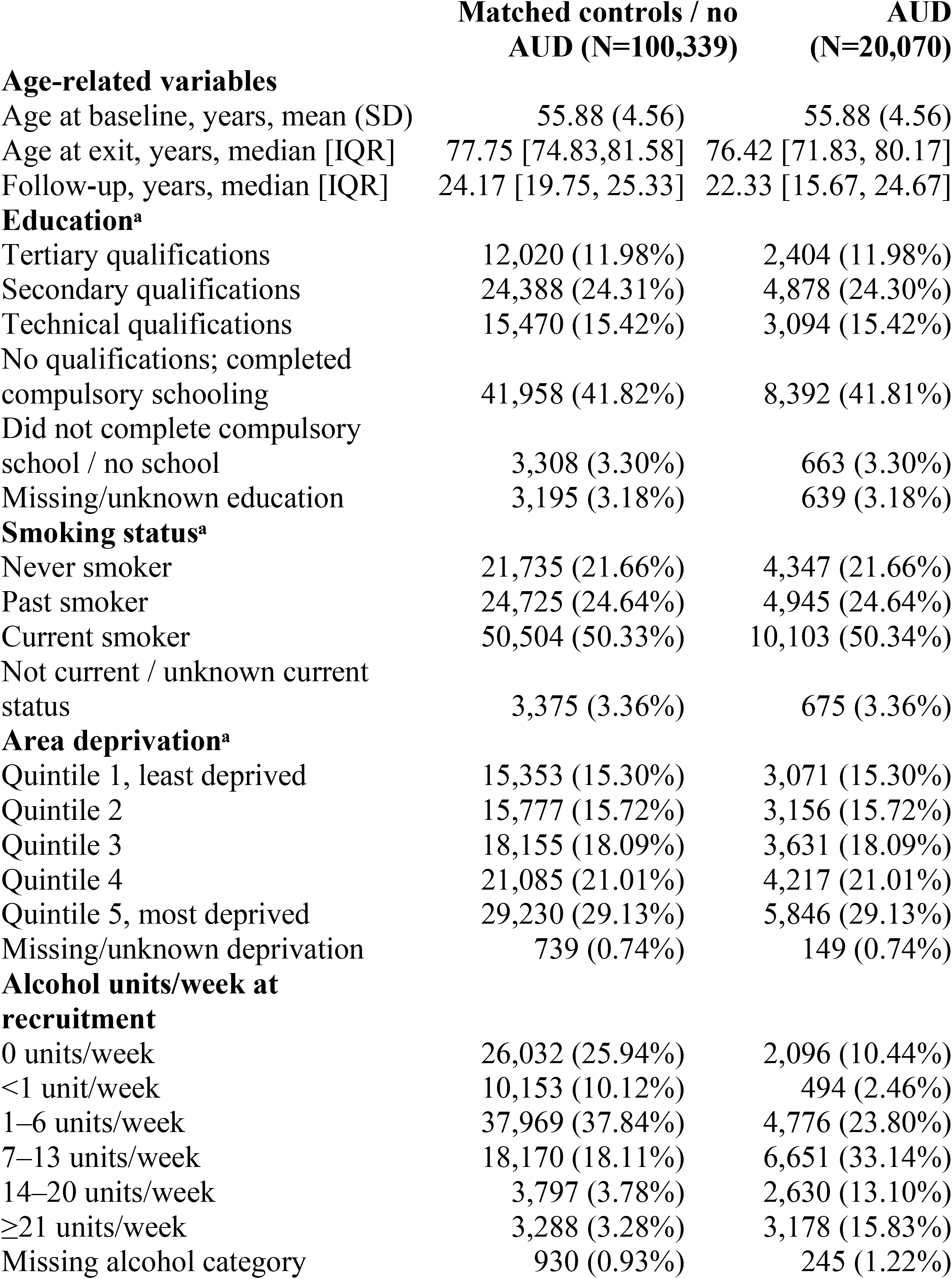
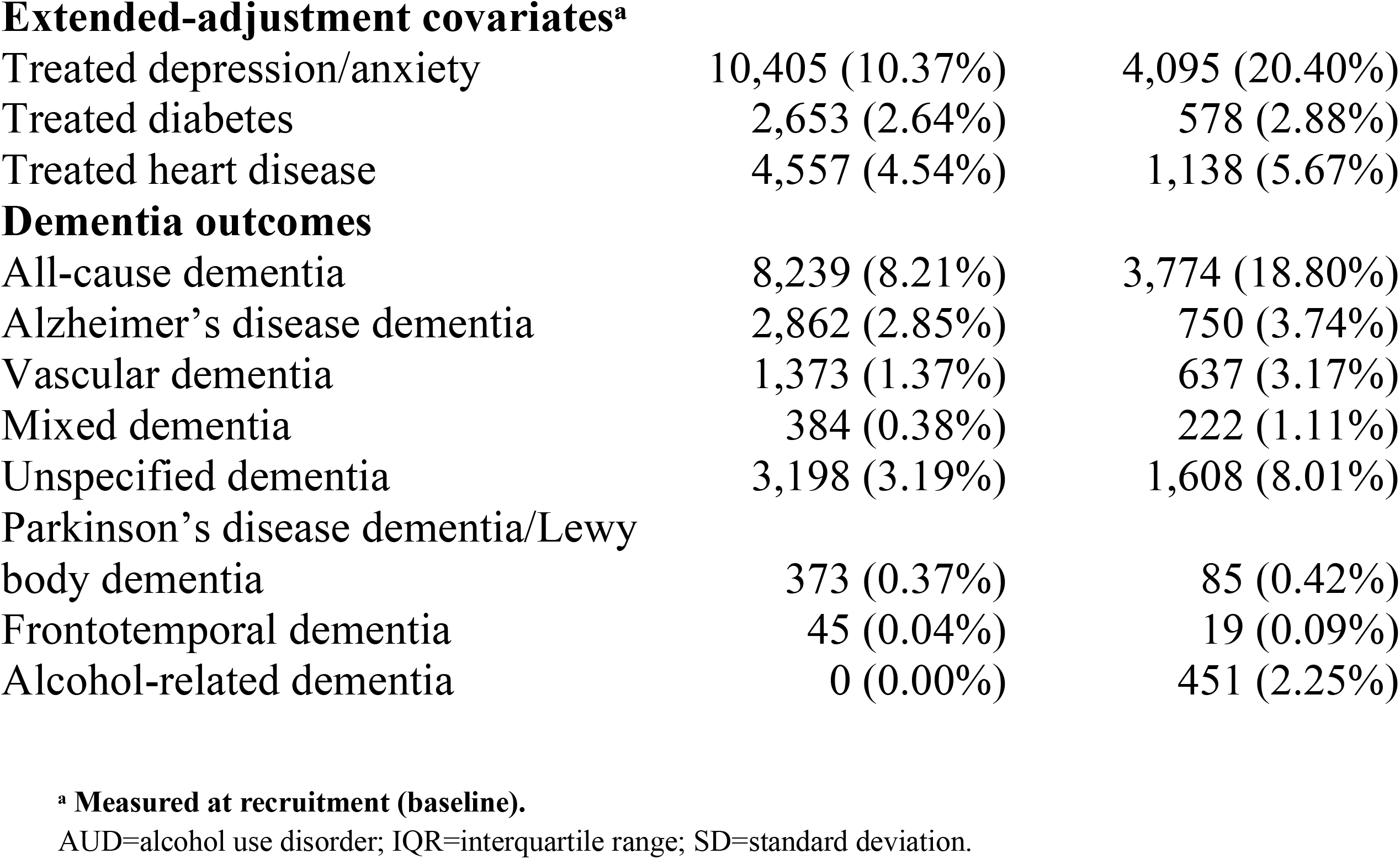

|  | Matched controls / no<br>AUD (N=100,339) | AUD<br>(N=20,070) |
| --- | --- | --- |
| **Age-related variables** |  |  |
| Age at baseline, years, mean (SD) | 55.88 (4.56) | 55.88 (4.56) |
| Age at exit, years, median [IQR] | 77.75 [74.83,81.58] | 76.42 [71.83, 80.17] |
| Follow-up, years, median [IQR] | 24.17 [19.75, 25.33] | 22.33 [15.67, 24.67] |
| **Education<sup>a</sup>** |  |  |
| Tertiary qualifications | 12,020 (11.98%) | 2,404 (11.98%) |
| Secondary qualifications | 24,388 (24.31%) | 4,878 (24.30%) |
| Technical qualifications | 15,470 (15.42%) | 3,094 (15.42%) |
| No qualifications; completed compulsory schooling | 41,958 (41.82%) | 8,392 (41.81%) |
| Did not complete compulsory school / no school | 3,308 (3.30%) | 663 (3.30%) |
| Missing/unknown education | 3,195 (3.18%) | 639 (3.18%) |
| **Smoking status<sup>a</sup>** |  |  |
| Never smoker | 21,735 (21.66%) | 4,347 (21.66%) |
| Past smoker | 24,725 (24.64%) | 4,945 (24.64%) |
| Current smoker | 50,504 (50.33%) | 10,103 (50.34%) |
| Not current / unknown current status | 3,375 (3.36%) | 675 (3.36%) |
| **Area deprivation<sup>a</sup>** |  |  |
| Quintile 1, least deprived | 15,353 (15.30%) | 3,071 (15.30%) |
| Quintile 2 | 15,777 (15.72%) | 3,156 (15.72%) |
| Quintile 3 | 18,155 (18.09%) | 3,631 (18.09%) |
| Quintile 4 | 21,085 (21.01%) | 4,217 (21.01%) |
| Quintile 5, most deprived | 29,230 (29.13%) | 5,846 (29.13%) |
| Missing/unknown deprivation | 739 (0.74%) | 149 (0.74%) |
| **Alcohol units/week at recruitment** |  |  |
| 0 units/week | 26,032 (25.94%) | 2,096 (10.44%) |
| <1 unit/week | 10,153 (10.12%) | 494 (2.46%) |
| 1–6 units/week | 37,969 (37.84%) | 4,776 (23.80%) |
| 7–13 units/week | 18,170 (18.11%) | 6,651 (33.14%) |
| 14–20 units/week | 3,797 (3.78%) | 2,630 (13.10%) |
| ≥21 units/week | 3,288 (3.28%) | 3,178 (15.83%) |
| Missing alcohol category | 930 (0.93%) | 245 (1.22%) |
| **Extended-adjustment covariates<sup>a</sup>** |  |  |
| Treated depression/anxiety | 10,405 (10.37%) | 4,095 (20.40%) |
| Treated diabetes | 2,653 (2.64%) | 578 (2.88%) |
| Treated heart disease | 4,557 (4.54%) | 1,138 (5.67%) |
| **Dementia outcomes** |  |  |
| All-cause dementia | 8,239 (8.21%) | 3,774 (18.80%) |
| Alzheimer's disease dementia | 2,862 (2.85%) | 750 (3.74%) |
| Vascular dementia | 1,373 (1.37%) | 637 (3.17%) |
| Mixed dementia | 384 (0.38%) | 222 (1.11%) |
| Unspecified dementia | 3,198 (3.19%) | 1,608 (8.01%) |
| Parkinson's disease dementia/Lewy<br>body dementia | 373 (0.37%) | 85 (0.42%) |
| Frontotemporal dementia | 45 (0.04%) | 19 (0.09%) |
| Alcohol-related dementia | 0 (0.00%) | 451 (2.25%) |

<sup>a</sup> Measured at recruitment (baseline).
AUD=alcohol use disorder; IQR=interquartile range; SD=standard deviation.

