## Supplementary Methods for "Clinically Recognised Alcohol Use Disorder and Risk of Dementia Subtypes in 1.3 Million Women: A Prospective Cohort Study"

#### **Exposure and outcome classification**

Self-reported alcohol consumption at recruitment was examined as a secondary exposure distinct from hospital-recorded AUD. The 0-units/week category included both never and former drinkers because these groups could not be distinguished from the recruitment questionnaire. Hospital-recorded AUD therefore represents a more severe, clinically recognised alcohol-related phenotype, whereas self-reported alcohol consumption reflects drinking behaviour reported at recruitment.

For each participant, all recorded dementia-related ICD-10 diagnoses were identified from Hospital Episode Statistics records and the date of each diagnosis was extracted. Dementia subtypes were assigned using a prespecified rule-based hierarchy developed with specialist clinical input. Participants with a single specific dementia subtype diagnosis were assigned that subtype. Participants with two or more specific dementia subtype diagnoses, such as Alzheimer’s disease and vascular dementia, were classified as having mixed dementia. Participants with an unspecified dementia diagnosis followed by a specific dementia subtype were assigned the subsequent specific subtype. In all cases, the date of the first recorded dementia diagnosis was retained as the dementia event date, even when the subtype classification was subsequently refined.

Mild cognitive impairment was identified as a separate cognitive phenotype and was not classified as dementia. Participants with mild cognitive impairment followed by dementia were assigned the subsequent dementia subtype, with the first qualifying dementia diagnosis used as the dementia event date. Participants with mild cognitive impairment only remained in the primary risk set and were excluded in a sensitivity analysis.

Non-dementia codes for Parkinson’s disease and Huntington’s disease were treated as marker codes rather than standalone dementia outcomes. These codes were considered supportive evidence for Parkinson’s disease dementia/Lewy body dementia and Huntington’s disease dementia, respectively, only when accompanied by a qualifying dementia code. Marker-only records without a qualifying dementia diagnosis were not classified as dementia cases.

**Alcohol-related dementia defined using ICD-10 codes F10.6 and G31.2, contributed to the all-cause dementia outcome but was not used to define AUD exposure. Because alcohol-related dementia overlaps diagnostically and conceptually with alcohol-related harm, it was not included among the primary subtype-specific Cox analyses. Participants with alcohol-related dementia but no qualifying AUD exposure were excluded from the non-AUD control pool, whereas AUD-exposed participants with alcohol-related dementia were retained. Full ICD-10 code lists for alcohol-related exposure and dementia outcomes are provided in the supplementary material and in machine-readable format in the study GitHub repository.**(<https://github.com/Topiwala-Lab/mws-aud-dementia>).

### **Statistical analyses**

#### Covariate and missing-data handling

Participants with missing baseline smoking status were excluded because smoking was a key matching variable and is strongly associated with AUD, vascular comorbidity, mortality, and dementia risk. Treating missing smoking status as a separate category could have resulted in participants being matched on smoking missingness rather than smoking behaviour.

Education, smoking status, area deprivation, and region were treated as categorical variables. Binary treatment and comorbidity variables were treated as yes/no indicators, and missing values were not retained as separate model categories. Cox models additionally adjusted for the matching variables to account for any residual measured imbalance after matching while preserving stratification by matched set.

#### Subtype-specific outcome models

For subtype-specific analyses, the target dementia subtype was treated as the event. Participants were censored at death, administrative end of follow-up, or diagnosis of another specific dementia subtype before the target subtype. Unspecified dementia and mild cognitive impairment were not treated as competing censoring subtypes in models of specific dementia subtypes.

#### Temporal exposure-lag analyses

To characterise how associations changed with increasing temporal separation between first recorded AUD and subsequent dementia, the time-varying AUD exposure was delayed by 0 to 15 years in annual increments.

For a lag of **L years**, a woman with AUD remained classified as unexposed until **L years** after her first qualifying AUD record and became exposed only after that delayed exposure date. Dementia occurring before the delayed exposure date was retained in the analysis and counted as an event during unexposed person-time. Participants were not excluded because dementia occurred within the lag interval.

The 0-year lag reproduced the primary time-varying AUD model. The same attained-age time scale, matched-set structure, endpoint-specific censoring rules, and Model 1 and Model 2 covariate sets were used across lag values.

These analyses therefore differed from conventional dementia washout or event-exclusion analyses. Their purpose was to characterise how the association changed as the required temporal separation between first recorded AUD and classification as exposed increased.

#### Mild cognitive impairment sensitivity analysis

To assess whether prodromal cognitive impairment without recorded dementia influenced the findings, the primary analyses were repeated after excluding women with mild cognitive impairment without a qualifying dementia diagnosis. Matched sets rendered incomplete by this exclusion were subsequently removed before the matched models were refitted.

#### Competing mortality and absolute cumulative incidence

In addition to the Fine–Gray and cause-specific mortality models described in the main Methods, absolute cumulative incidences of dementia and death before dementia were estimated using the Aalen–Johansen estimator. For each matched set, follow-up began at the attained age of the AUD participant’s first recorded AUD diagnosis, or at recruitment if AUD was already recorded at or before baseline; matched controls entered at the same attained age provided they remained under observation, alive, and dementia-free. Dementia and death before dementia were treated as competing events, and cumulative incidence was summarised at 5 and 10 years after this matched index point.

#### Calendar period adjustment

To examine whether secular changes in hospital coding, diagnostic practice, or dementia ascertainment influenced the results, follow-up was split into calendar periods of 1996–2004, 2005–2009, 2010–2014, 2015–2018, and 2019 onwards. Calendar period was entered as a time-varying covariate in Cox models with attained age as the underlying time scale and matched set as the stratification variable. AUD remained time-varying and the models otherwise used the same adjustment sets as the primary analyses. These estimates represent overall AUD hazard ratios adjusted for calendar period rather than calendar period-specific hazard ratios.

#### Baseline alcohol consumption by attained age

As an extension of the baseline alcohol consumption analysis described in the main Methods, associations were additionally estimated within attained-age bands of <70, 70–79, and ≥80 years using the same alcohol-consumption categories, reference group, and covariate adjustment.

#### Software

Data management was performed primarily using the data.table package in R. Cox proportional hazards models were fitted using the survival package, and Fine–Gray analyses were implemented using finegray() and coxph(). Figures were generated using ggplot2, with grid and gridExtra used for graphical and tabular outputs.

**Supplementary figure captions:**

**Supplementary Figure 1. Associations between baseline self-reported alcohol consumption and all-cause dementia by attained-age band.** Hazard ratios (HRs) and 95% confidence intervals (CIs) for incident all-cause dementia according to self-reported alcohol consumption at recruitment, estimated separately within attained-age bands of <70, 70–79, and ≥80 years. The reference category was 1–6 units/week. Estimates are from Model 2 and were adjusted for education, smoking status, area deprivation, region, treated depression/anxiety, diabetes, and cardiovascular disease. Baseline alcohol consumption was treated as a fixed exposure.

**Supplementary Figure 2. Comparison of primary AUD associations with mild cognitive impairment exclusion and calendar-period-adjusted analyses.** Hazard ratios (HRs) and 95% confidence intervals (CIs) for the association between time-varying hospital-recorded alcohol use disorder (AUD)/alcohol-related harm and incident all-cause dementia and dementia subtypes. Primary Model 2 estimates are shown alongside estimates obtained after excluding women with mild cognitive impairment without a qualifying dementia diagnosis and estimates additionally adjusted for calendar period as a time-varying covariate. All models used attained age as the underlying time scale, retained the matched-set structure, and adjusted for education, smoking status, area deprivation, region, treated depression/anxiety, diabetes, and cardiovascular disease. Calendar-period-adjusted estimates represent overall AUD associations adjusted for secular period rather than period-specific hazard ratios.
